# MCH-Guard: Multimodal Machine Learning Framework for Risk Stratification of Cerebral Microhemorrhage Risk in the Alzheimer’s Disease Neuroimaging Initiative

**DOI:** 10.64898/2026.06.18.26355972

**Authors:** Alper Gel, Eliana Phillips, Isabella Hausle, Pamela Thropp, Duygu Tosun, Alzheimer’s Disease Neuroimaging Initiative

## Abstract

**Background:** Efficient cerebral microhemorrhage (MCH) monitoring is critical for anti-amyloid therapy safety due to ARIA-H risk. We developed MCH-Guard, a multimodal machine-learning framework, to stratify MCH risk using ADNI data (N=813).

**Methods:** Nested models integrated clinical history, fluid biomarkers, and imaging to predict MCH presence, incidence, and stability.

**Results:** The comprehensive model detected baseline MCH with high accuracy (AUC 0.86). Notably, the “minimal” model (M1), utilizing only demographics and clinical history, achieved robust performance (AUC 0.82). Longitudinal models predicted time-to-onset (R^2^=0.68) and stratified four-year risk. Furthermore, we identified a transient vascular instability phenotype—where MCH status fluctuates— which was strongly predicted by hepatic factors.

**Conclusions:** MCH-Guard offers a flexible clinical decision-support tool for optimizing spontaneous MCH & ARIA surveillance. The strong performance of the clinical-only M1 model supports equitable risk assessment in resource-limited settings, while the characterization of vascular instability addresses a critical confounder in safety monitoring.

## 1. BACKGROUND

Cerebral microhemorrhages (MCH) are small-vessel bleeds detectable on T2*-weighted magnetic resonance imaging (MRI) as hypointense lesions. They are common in older adults, utilized as a neuroimaging marker of cerebral small vessel disease[1,2] and associated with cognitive decline, increased risk of stroke, and mortality[3–6]. A primary pathological substrate for MCH—especially in lobar regions (i.e., grey-white matter junction)—is cerebral amyloid angiopathy (CAA). CAA is characterized by the deposition of amyloid-β (Aβ) peptides within the walls of cerebral arteries and arterioles, a condition present in up to 90% of individuals with Alzheimer’s disease (AD)[7]. CAA compromises vascular integrity, leading to vessel wall fragility and a propensity for rupture, which manifests as MCH and, in more severe cases, intracerebral hemorrhage[8]. Thus, MCH is considered a principal in-vivo marker of underlying CAA pathology.

The vascular fragility associated with CAA has become a central challenge in the era of amyloid-targeting therapies (ATTs) for AD. These treatments can provoke inflammatory responses and increase vascular permeability, leading to a spectrum of MRI findings known as Amyloid-Related Imaging Abnormalities (ARIA)[9]. ARIA is classified as two types: ARIA with edema or effusions (ARIA-E) and ARIA with hemosiderin deposition (ARIA-H), which includes new MCH and superficial siderosis[10]. Crucially, the presence of baseline MCH[11,12] and *APOE* ε4 allele[13,14] are the strongest risk factors of developing ARIA during ATT treatments.

This heightened risk has led to stringent eligibility criteria for these therapies, often excluding individuals with more than four MCH from clinical trials[15] and requiring frequent, resource-intensive MRI monitoring to detect ARIA-H. Current FDA and EMA guidelines mandate rigorous surveillance schedules that strain healthcare systems and place a burden on patients. There is an urgent unmet need for data-driven tools capable of triaging patients into risk categories. While distinct, spontaneous MCH and ARIA-H share a common substrate—vascular amyloidosis—making spontaneous risk a viable proxy for therapeutic safety stratification in the absence of trial data. A precision prognostic engine that identifies high-risk individuals could optimize resource allocation, ensuring that intensive surveillance is targeted toward those most vulnerable to vascular events, while potentially reducing the burden on lower-risk populations.

A growing body of evidence has identified key factors associated with MCH. Among the most established predictors are advancing age and chronic hypertension, linked particularly to MCH in deep brain regions[16,17]. The *APOE* ε4 allele is strongly associated with the presence of strictly lobar MCH, underscoring the shared pathological association with CAA[18–20]. Further, lower cerebrospinal fluid (CSF) levels of both Aβ40 and Aβ42 have been associated to a higher burden of lobar MCH, reflecting the sequestration of Aβ within the cerebral vessel walls in CAA[21–24]. Similarly, lower CSF p-tau181 levels are negatively correlated with higher lobar MCH burden[25]. Concurrently, elevated plasma and CSF levels of neurofilament light chain (NfL), a marker of neuroaxonal injury, have been associated with the overall burden of cerebral small vessel disease, including MCH[26,27]. Imaging studies have also shown strong associations between MCH and reduced hippocampal volume with increased white matter hyperintensities[10,21,21,28–30].

Much of this evidence comes from cross-sectional studies identifying factors associated with MCH prevalence. A critical knowledge gap persists in understanding the predictors of MCH incidence, either spontaneous or due to ATTs. Identifying the clinical, genetic, and biomarker profiles of individuals who are actively developing MCH is essential for risk stratification. Therefore, the primary objective of this study was to identify factors associated with both the presence of MCH at baseline and the incidence of new MCH over a follow-up period in a large observational cohort of Alzheimer’s Disease Neuroimaging Initiative (ADNI), where 36% of participants presented with spontaneous MCH at baseline. In this study, we introduce MCH-Guard, a multimodal machine-learning framework integrating patient-level imaging, biomarkers, and clinical history for personalized MCH risk prediction. To ensure clinical utility, MCH-Guard uses a series of nested models that incrementally incorporate features, allowing flexible application across settings with varying data availability.

## 2. METHODS

### 2.1 Study design and participants

Data was obtained from the Alzheimer’s Disease Neuroimaging Initiative (ADNI) database (https://adni.loni.usc.edu/). The ADNI study was launched in 2004 as a public-private partnership. The primary goal of ADNI has been to test whether serial magnetic resonance imaging (MRI), positron emission tomography (PET), other biological markers, and clinical and neuropsychological assessment can be combined to measure the progression of mild cognitive impairment (MCI) and early AD. Cognitively unimpaired (CU) individuals, individuals with MCI, and individuals clinically diagnosed with dementia due to AD from the ADNI GO/2/3 phases were included in this current study. Within those cohorts, participant inclusion was based on whether subjects had available MRI MCH assessments, medical history questionnaire data, information on their concurrent medications, demographic information, plasma NfL data, *APOE* ε4 genotype, CSF biomarkers of Aβ42 and p-tau181, and MRI data for assessment of cerebrovascular integrity and anatomical phenotypes. This multi-modal data flow is illustrated in Supplemental Figure S1.

### 2.2 Data Modalities

#### 2.2.1 MCH assessment on T2*-weighted MRI

T2*-weighted gradient echo MRI scans (3mm axial, 1mm in-plane resolution) from the ADNI GO/2/3 cohorts were used to identify MCH by the ADNI MRI Core [31]. MCH, superficial siderosis, and “other” findings were annotated as “observations” with location data in the native coordinate system of each T2* MRI. Briefly, for each of the N available T2* MRIs per participant, the other N-1 images were affine-registered and resampled into the current image space. This allowed for visual comparison of findings across scans, but assessment and annotation were performed only on the current image. For each T1-weighted image (typically from the same session), tissue probability maps (SPM5) and a 35-region atlas were generated.

The T1 image, along with the atlas (nearest neighbor interpolation) and probability maps (trilinear interpolation), were registered and resampled into the current T2* MRI space. Image intensities were linearly scaled to standardize white matter. Analysts reviewed each T2* MRI, with findings identified in previous timepoints displayed on their respective MRIs. Findings could be “brought forward” to the current image for review and annotation. The final output consisted of a list of findings, each with one or more observations. Each observation included location data, MCH status (possible, definite, rescinded), tissue probability estimates, and atlas region assignment. In the current study, a participant’s scan was classified as MCH positive (MCH-pos) if the T2* reading at that time indicated the presence of any MCH.

#### 2.2.2 Medical History

Initial medical history assessment during each individual’s screening visit included the presence (or absence) of the following comorbid health conditions: 1. Psychiatric; 2. Neurologic (other than AD); 3. Head, Eyes, Ears, Nose and Throat; 4. Cardiovascular; 5. Respiratory; 6. Hepatic; 7. Dermatologic-Connective Tissue; 8. Musculoskeletal; 9. Endocrine-Metabolic; 10. Gastrointestinal; 11. Hematopoietic lymphatic; 12. Renal-Genitourinary; 13. Allergies or Drug Sensitivities; 14. Alcohol Abuse; 15. Drug Abuse; 16. Smoking; 17. Malignancy; 18. Major Surgical Procedures; 19. Other.

#### 2.2.3 Concurrent Medications

Longitudinal medication data, collected in free-text format at each participant’s visit throughout the study period, was processed to create a structured, quantitative dataset. We used Chat-GPT 4o, a Large Language Model (LLM) and a taxonomy-based encoding system to convert the unstructured free-text strings into a hierarchical classification [32]. Specifically, hierarchical taxonomy of medication classes was established. This taxonomy was structured with multiple levels to capture increasing specificity. For example, a medication might be classified at the top level (general class) as an “Antidepressant”, at the second level (subclass) as a “Selective Serotonin Reuptake Inhibitor (SSRI)”, and at the third level (sub-subclass) as a specific compound like “Sertraline”. The full taxonomy used is detailed in Supplemental Table S1. The LLM was prompted to map each free-text medication string to the most appropriate level within the defined taxonomy, grounding the result with an LLM web-search tool to ensure realistic medication mapping. The model was given specific instructions to handle variations in spelling, common abbreviations, and different drug names (brand versus generic) by aligning them with the corresponding medication class in our taxonomy. The output of this process was a longitudinal data table encoded numerical categories representing the medication’s general class, subclass, and sub-subclass, according to the established taxonomy. The MCH-Guard models utilized the medication subclass feature. Based on the taxonomy at time of publication, the cohort has 96% of medication data entries as a classified item in the taxonomy, with the remaining being classified as ‘other’.

#### 2.2.4 Participant Demographics

Demographic data included age (in years), self-reported highest educational attainment (greater than or equal to college educated or not), self-reported sex (male, female), self-reported ethnoracial background (Non-Hispanic White [NHW], Non-Hispanic Black [NHB], Hispanic/Latin American [HLA], and other).

#### 2.2.5 Clinical Assessments

MCH-Guard utilizes a combination of harmonized cognitive and cardiovascular data from the Alzheimer’s Disease Sequencing Project (ADSP) Phenotype Harmonization Consortium (PHC), along with diagnostic and cognitive impairment severity measures. The ADSP-PHC datasets were generated through the large-scale harmonization of cardiovascular and cognitive measures across multiple AD cohorts, including ADNI, NACC, ROS/MAP, and WHICAP (https://adsp.niagads.org/). Cardiovascular variables (e.g., hypertension, diabetes, stroke, body mass index (BMI)) were standardized from self-reported and measured health variables to create composite scores. Participants underwent comprehensive clinical assessments, including collateral interviews, neurological examinations, and the Clinical Dementia Rating® (CDR®) and the CDR-Sum of Boxes (CDR-SB). Neuropsychological test items were quality-checked, anchored, and aligned across studies using psychometric modeling. This co-calibration produced harmonized, domain-specific scores (memory, executive functioning, language) that are directly comparable despite differences in the original test batteries [33].

#### 2.2.6 CSF AD Biomarkers

CSF Aβ42 and phosphorylated-tau181 (p-tau181) levels were measured in singlicate using Roche Elecsys ECLIA assays on a cobase 601 analyzer at the ADNI Biomarker Laboratory, following manufacturer instructions and a Roche study protocol [34]. A single reagent lot was used for each biomarker. Analyses were performed between November 17, 2016, and June 22, 2022, using a standard new lot rollover protocol with quality control samples. P-tau181/Aβ42 ratio was calculated as an overall measure of AD pathology [35].

#### 2.2.7 Plasma NfL

Plasma NfL was analyzed by the Single Molecule array (Simoa) technique by the Clinical Neurochemistry Laboratory at University of Gothenburg, Sweden [36]. The assay uses a combination of monoclonal antibodies, and purified bovine NfL as a calibrator. All samples were measured in duplicate, except for one (due to technical reasons). Analytical sensitivity was less than 1.0 pg/mL, and no sample contained NFL levels in plasma below the limit of detection (LOD).

### 2.2.8 MRI Volumetrics: White Matter Hyperintensities and Gray Matter

White matter hyperintensity (WMH) segmentation used a Bayesian approach combining 3D T1 and FLAIR MRI [37]. First, non-brain tissue was removed from T1 images using an atlas-based method. FLAIR images were affine registered to the T1s (FLIRT). T1 inhomogeneity was corrected via interleaved bias estimation and Bspline deformation, while FLAIR inhomogeneity was corrected using local histogram normalization after co-registration. T1 images were then non-linearly aligned to a standard template, and the corresponding FLAIRs were transformed to this space. WMH probability maps were generated from more than 700 individuals with semi-automatic WMH detection and manual editing. WMH likelihood was estimated through histogram segmentation and thresholding. Bayesian segmentation, combining image likelihood, spatial priors, and tissue constraints, was performed in standard space. Resulting WMH probability maps were thresholded (3.5 SD above the mean) to create binary masks. These masks were then inversely transformed into native space for WMH volume calculation. Gray matter volumes were obtained from T1-weighted MRI scans using automated tissue segmentation methods. After preprocessing steps (including skull stripping, bias field correction, and registration to a common template), gray matter, white matter, and cerebrospinal fluid were segmented using probabilistic tissue classification algorithms. Total gray matter volume was then calculated by summing all voxels classified as gray matter within the cerebrum. To account for individual differences in head size, both WMH and total gray matter volume were normalized to total intracranial volume (ICV).

### 2.2.9 T1 MRI Latent Embeddings

Structural T1-weighted MRIs were used to develop a quantitative imaging phenotype. To accomplish this, we first generated a high-dimensional latent space representation of the T1 images. This was achieved using a self-supervised, 3D convolutional autoencoder (CAE) model, which had been preliminarily developed and trained on a large, diverse dataset from the UK Biobank [38]. The CAE’s objective is to learn a compressed representation of the input data, often referred to as a latent embedding, which captures the salient features of the input T1 images in a more compact form. The pre-trained model was then fine-tuned on the ADNI dataset to optimize its performance specifically for the characteristics of the ADNI T1-weighted MRIs. The model’s performance was evaluated on a held-out test set of unseen data from the ADNI cohort. Details are provided in Supplementary Method S1. Following this, each T1-weighted image was encoded into a 128-dimensional latent vector, representative of an Unsupervised Deep learning derived Imaging Phenotypes (UDIP) space. To facilitate downstream analysis and respect inherent non-linearity of UDIP space, we applied Locally Linear Embedding (LLE) to reduce the dimensionality of the UDIP space [39]. The 128-dimensional UDIP representations were then projected to the 1st phenotypic axis of LLE, representing the most prominent, continuous anatomical variation captured within each 128-dimensional UDIP space. In a post-hoc analysis, high scores on The 1st Phenotypic Axis correlated strongly with patterns of ventricular enlargement and temporal atrophy, providing a nonlinear composite metric of neurodegenerative severity.

### 2.3. Statistical Analyses and Modeling

Statistical analysis was conducted in Python using the NumPy, Scikit-Learn, and Optuna libraries. All statistical tests were two-sided. For each prediction task, we constructed nested models in which each successive model added additional feature domains (M1⊂M2⊂M3). Pairwise differences in model performance metrics were evaluated using McNemar’s test for classification tasks and permutation tests for regression tasks, with 95% bootstrap confidence intervals reported where indicated.

#### 2.3.1 Prediction of MCH Presence (Classification Models)

To predict the presence of MCH at a single time point, we trained Random Forest classification models (CLS) to distinguish MCH-positive (MCH-pos) from MCH-negative (MCH-neg) individuals. The nested CLS models used the following feature sets: CLS-M1 included demographics, *APOE* ε4 genotype, comorbid conditions, concurrent medication use history, cognitive assessments; CLS-M2 included CLS-M1 features plus plasma NfL and CSF p-tau181/Aβ42 ratio; CLS-M3 included CLS-M2 features plus ICV-adjusted WMH burden, ICV-adjusted total gray matter volume, and the 1^st^ phenotypic axis from T1-weighted MRI latent embedding. Data were split into training, validation, and test sets using an 80%/10%/10% stratified split. Continuous features were Z-score normalized using scikit-learn’s StandardScaler. Hyperparameters were tuned with Optuna using 100 trials (CLS-M1), 200 trials (CLS-M2), and 250 trials (CLS-M3), where trial count is scaled by the number of additional features added.

Hyperparameter tuning utilized a median-pruner strategy to maximize validation accuracy. Final models were refitted on the full training set with optimized hyperparameters. Performance was evaluated on the test set using ROC-AUC and accuracy, with 95% bootstrap confidence intervals. Feature importance was estimated using mean decrease in impurity (MDI).

### 2.3.2 Prediction of Incidence of MCH Over Time (Longitudinal Models)

For prediction of incidence of MCH over time, we analyzed participants devoid of MCH at baseline with at least one follow-up MCH assessment. Time-to-event for incident MCH positivity was computed in years from baseline. Two longitudinal model families were evaluated: an ExtraTrees regressor (RG) to predict time to MCH positivity, and a L1-penalized Cox proportional hazards (CoxPH) survival model (SRV) to estimate patient-specific MCH risk curve over the next 48 months, offering insight into the change in MCH risk over extended periods. Prior to model fitting, predictors with high pairwise correlation (i.e., |ρ| > 0.9) or near-zero variance (i.e., σ < 0.01) were removed to facilitate model convergence. Specifically, sex, ethnoracial background, injectable diabetes medication usage, miscellaneous respiratory medication usage, Parkinson’s medication usage, and anti-arrythmia medication usage were removed due to having low variance.

The RG models were trained using a stratified 80%/20% train-test split. ExtraTreesRegressor hyperparameters were not tuned, as preliminary hyperparameter tuning experiments using RandomizedSearchCV resulted in overfitting; therefore, default hyperparameters were used. Model performance was evaluated on the test set using R^2^, mean squared error (MSE), root mean squared error (RMSE), and mean absolute error (MAE). Statistical comparisons among RG models were performed using permutation tests on R^2^ and RMSE.

For the SRV models, only baseline biomarker and imaging values were used as predictors. The SRV models were trained using an 80%/20% stratified train-test split. Hyperparameters were optimized with Optuna over 50 trials, tuning the elastic net penalization parameters (penalizer: 10 –10 on a log scale; l1 ratio: 0–1). A 10-fold cross-validation scheme was used within the training data to maximize the concordance index (C-index). The optimal penalized CoxPH model was then refitted on the full dataset with the selected hyperparameters. Model evaluation reports the C-index, partial Akaike information criterion (pAIC), and coefficient estimates with p-values. For each participant, the fitted model produces a continuous risk score representing the relative hazard of incident MCH positivity, which are then used to stratify patients into three risk groups using tertile-based discretization. Higher scores indicate greater risk of the adverse effect occurring sooner. While the Cox-PH model generates a continuous hazard ratio, clinical decision-making often requires categorical thresholds to triage patient care. To evaluate the model’s translational utility, we stratified participants into three distinct risk tiers (’Low,’ ‘Medium,’ and ‘High’) using quantile-based discretization of the predicted risk scores. This stratification allows us to assess the model’s ability to distinguish between individuals likely to remain MCH-free and those requiring heightened surveillance.

Finally, an exploratory analysis was conducted in a sub-cohort comprising approximately 15% of the longitudinal cohort, exhibiting at least one transition between one state to another and back (e.g., MCH-pos to MCH-neg then to MCH-pos) across follow-up visits. An XGBoost classifier (SW) was trained to predict patient assignment into this “switching” behavior sub-cohort. Statistical significance for classifier comparisons across the SW model set was assessed using McNemar’s test, and 95% bootstrap confidence intervals were reported for performance metrics.

## 3. RESULTS

### 3.1 Cohort Characteristics

A total of 813 unique individuals with 1706 datapoints were included in the final dataset, with 28.6% classified as MCH positive (MCH-pos) at baseline. Clinical and biomarker characteristics for MCH-pos and MCH negative (MCH-neg) groups are presented in Table 1. Compared to MCH-neg participants, those with MCH at baseline exhibited significantly different profiles across several domains. Specifically, they were significantly older (MCH-pos: 74.2 ± 7.0 vs MCH-neg: 71.3 ± 7.3, p<0.001), had a greater proportion of males (MCH-pos: 59.1% vs MCH-neg: 50.2%, p=0.027), and *APOE* ε4 homozygotes (MCH-pos: 12.9% vs MCH-neg: 8.1%, p=0.046). MCH-pos individuals showed significantly lower cognitive performance across memory (p < 0.001), executive function (p=0.005), and language domains (p = 0.0038), and more severe cognitive impairment (CDR-SB, p = 0.0025; Diagnosis, p=0.0115). They had a greater burden of CSF p-tau181/Aβ42 ratio (p < 0.001), normalized white matter hyperintensities (p < 0.001) and plasma NfL (p < 0.001). They also presented with lower BMI (p=0.0137), a greater frequency of cardiac (p=0.0027) and renal conditions (p=0.012), and increased use of AD and dementia-related medications (p=0.036) classified under the medication encoding taxonomy (Supplementary Table S1).

**Table 1.**
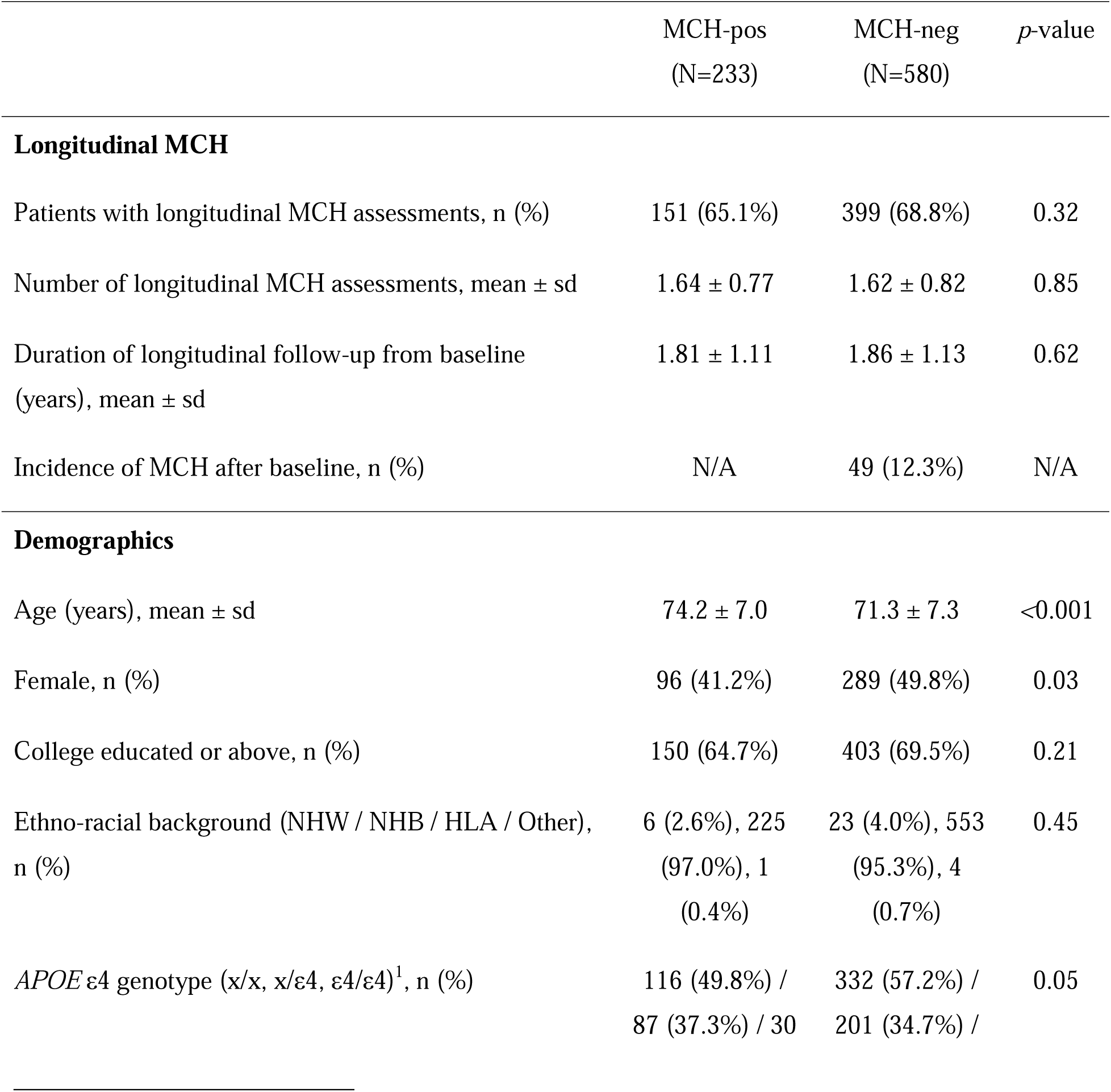
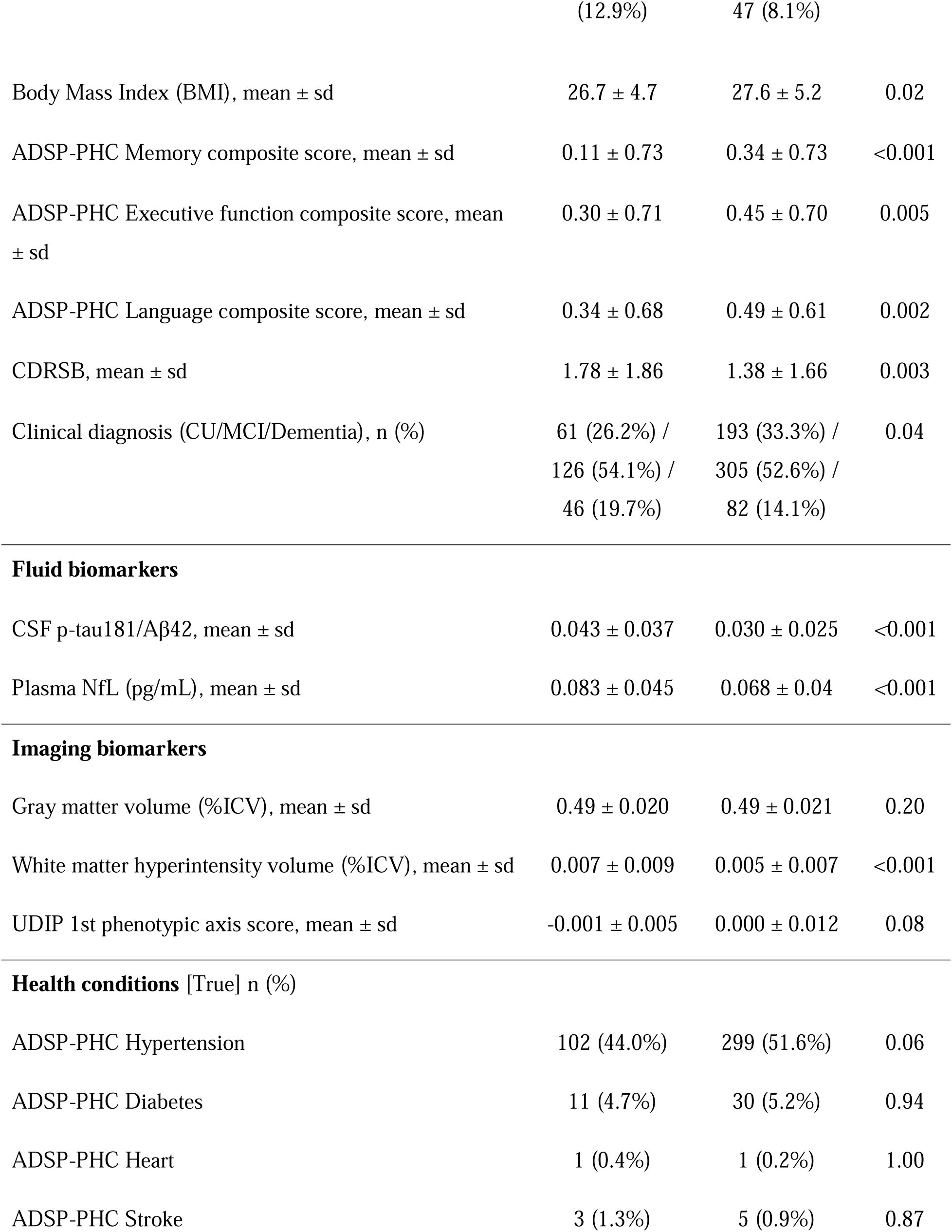
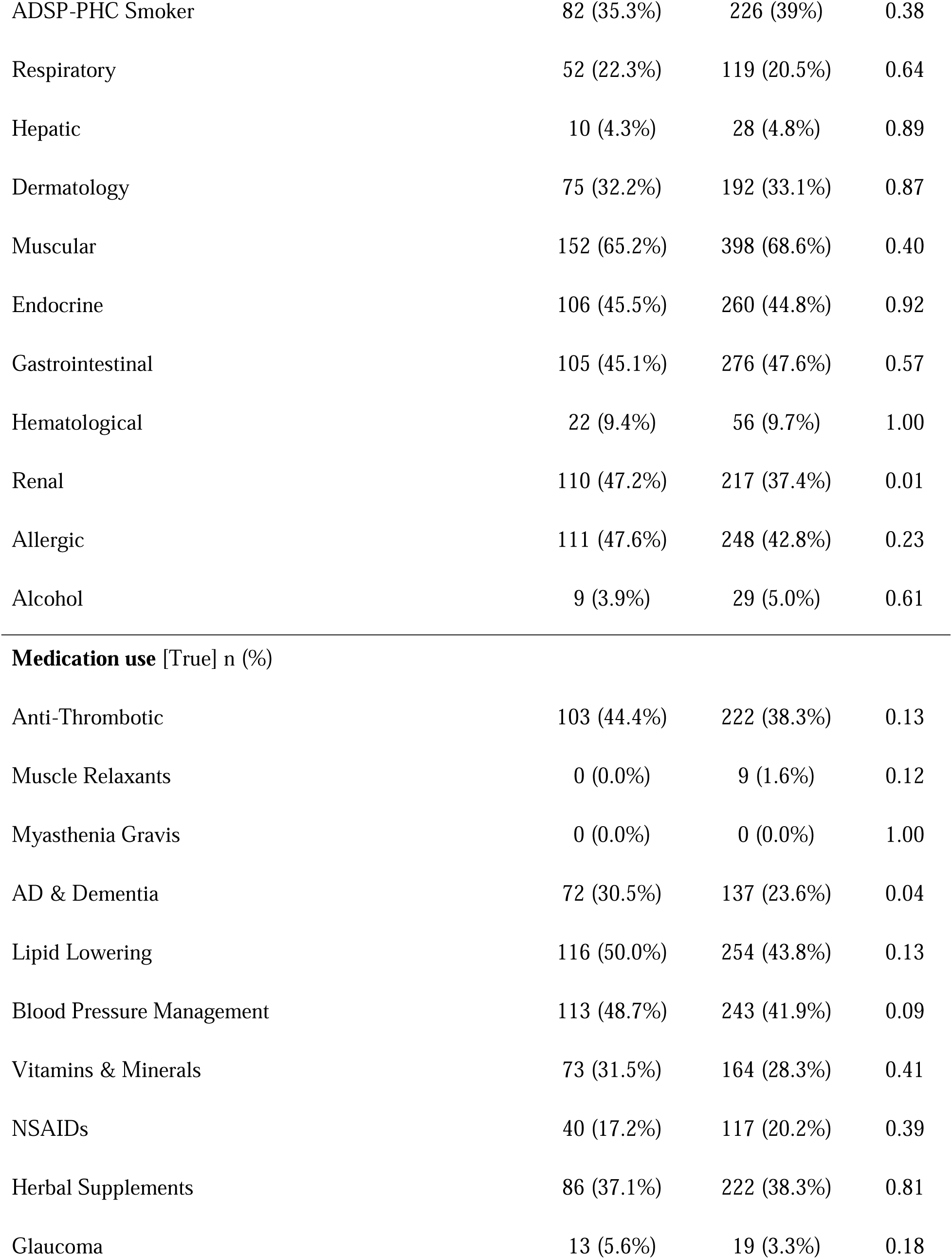
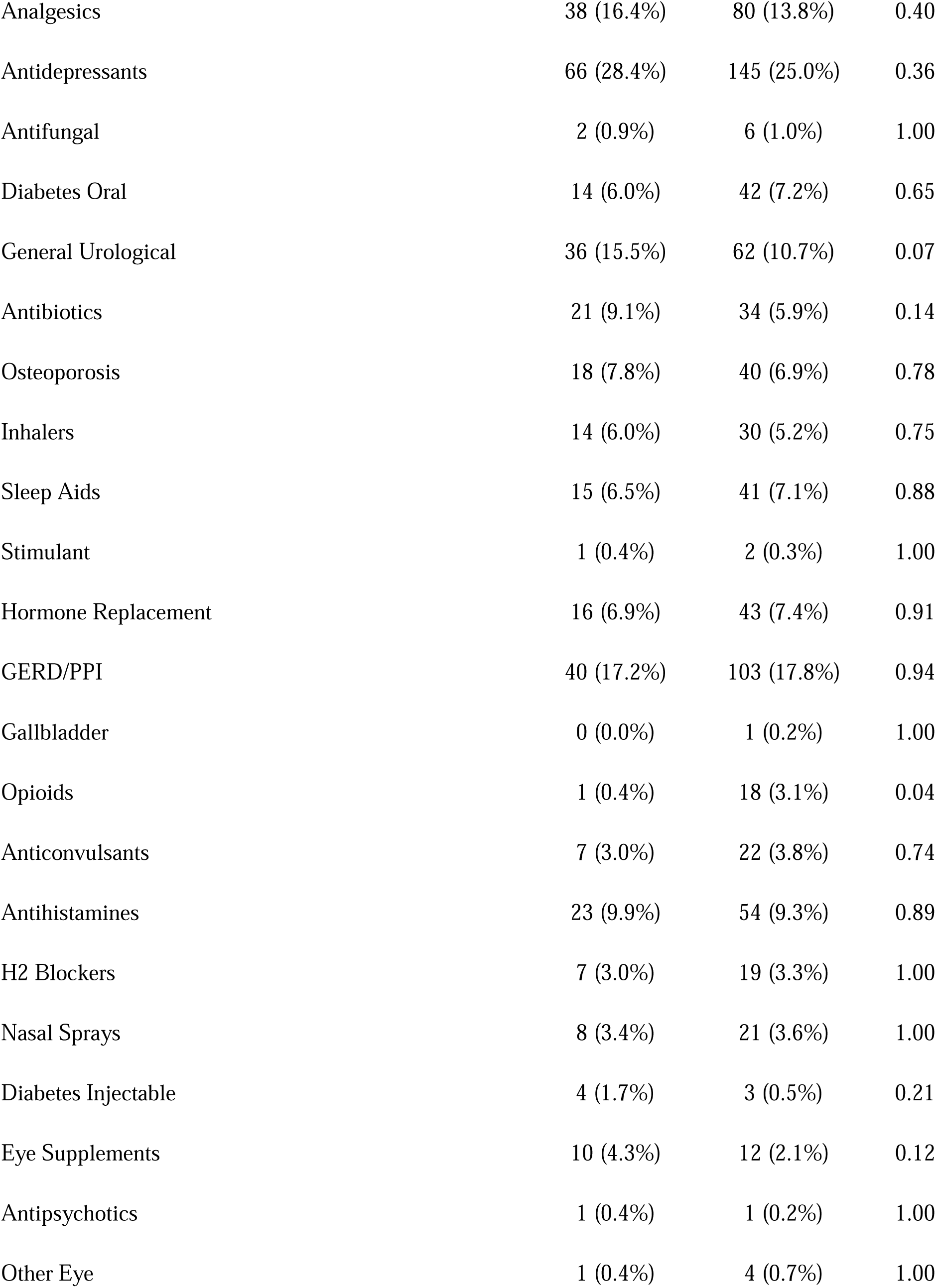
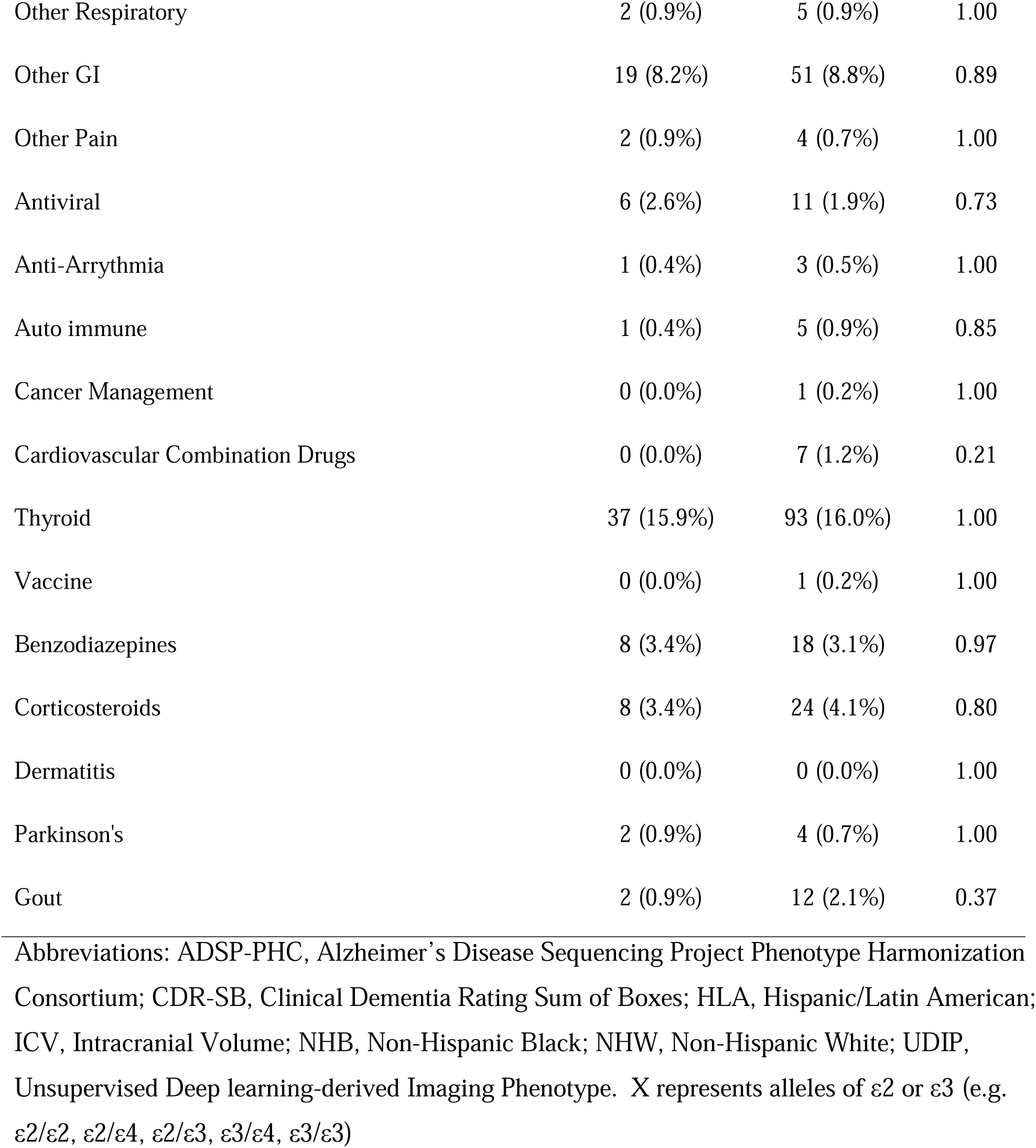
Baseline Demographic, Clinical, and Biomarker Characteristics of the Study Cohort Stratified by Cerebral Microhemorrhage (MCH) Status. Data are presented as mean ± standard deviation (SD) for continuous variables and count (percentage) for categorical variables. The cohort (N=813) is stratified into participants with MCH (MCH-pos) and without MCH (MCH-neg) at baseline. Statistical comparisons were performed using Fisher’s test for longitudinal variables, independent t-tests for continuous variables, and chi-squared tests for categorical variables.

Among those who were MCH-neg at baseline, 68.8% (n=399) had follow-up MCH assessment data and 11.5% (n=46) had developed MCH in their study follow-up of up to 1.86 ± 1.13 years. The majority of MCH-neg patients at baseline remained MCH-neg at their last follow-up or had no follow-up data (Supplementary Figure S2). Of those who were MCH-pos at baseline, 65.1% (n=151) had follow-up MCH assessment data, and only 4% (n=6) of those improved to MCH-neg at their last observation, with the remainder acquiring more MCH sites or remaining stable (Supplementary Figure S2).

### 3.2 Predictors of presence of MCH [CLS models]

To predict the presence of MCH at a given time point, we developed three nested classification (CLS) models. The performance of these models improved with the addition of more complex data modalities. Specifically, the CLS-M1 model, utilizing only demographic and clinical data, achieved a test-set accuracy of 77.2% ± 4.7% and an AUC of 0.82 ± 0.049 (Table 2; Figures 1a-b). The most important predictive features for this model were patient BMI, age and composite scores of memory, executive function, and language (Figure 1e). The CLS-M2 model, which added fluid biomarkers to the CLS-M1 features, yielded an improvement with an accuracy of 81.3% ± 4.4% with an AUC of 0.83 ± 0.053. In this variant, the CSF p-tau/Aβ42 ratio and plasma NfL levels emerged as key predictors, followed by the same top-features observed with CLS-M1 (Figure 1d). The CLS-M3 model, which incorporated imaging biomarkers, was the best-performing variant, achieving a test-set accuracy of 81.6%±4.7% and an AUC of 0.85 ± 0.051. The improvement in performance of CLS-M3 and CLS-M2 versus CLS-M1 was statistically significant shown via McNemar’s test (M3-M1: p = 0.032, M2-M1: p = 0.024). The most influential features for M3 were the p-tau/Aβ42 ratio, ICV-normalized white-matter-hyperintensities, BMI, plasma NfL levels, and 1 phenotypic axis score (Figure 1c). Complete performance metrics for all CLS model variants are detailed in Table 2.

**Figure 1:** Cross-Sectional Prediction of MCH Presence (Classification Models). **(A)** Receiver Operating Characteristic (ROC) curves comparing the performance of the nested model variants: M1 (blue), M2 (green), and M3 (red). **(B)** Comprehensive performance metrics (Accuracy, Precision, Recall, F1-Score, ROC-AUC) for all three models on the independent test set. Error bars represent 95% confidence intervals. **(C–E)** Signed feature importance plots for the M3, M2, M1 models, respectively. Bars indicate the magnitude of the feature’s contribution to the model, while color denotes the directionality of the effect (Red = Positive effect/Increased Risk; Green = Negative effect/Decreased Risk).

**Table 2.** Performance Metrics of the MCH-Guard Nested Machine Learning Models. Quantitative evaluation of the three model variants (M1: Clinical/Demographics; M2: +Fluid Biomarkers; M3: +Imaging Biomarkers) across four prediction tasks. Classification (CLS) models predict the cross-sectional presence of MCH, reported via Accuracy and Area Under the Curve (AUC). Regression (RG) models predict time-to-incidence of MCH in years, reported via R^2^, Root Mean Squared Error (RMSE), and Mean Absolute Error (MAE). Survival (SRV) models estimate the risk of MCH onset over time, evaluated by the Concordance Index (C-index) and Partial Akaike Information Criterion (pAIC). Switching (SW) models predict unstable MCH status. Values represent performance on the withheld test set with 95% confidence intervals where applicable. Bold values indicate the best-performing model for each task. **Bold** values indicate the best-performing model for each task. Arrows for each metric indicate desired direction for best performance. All values are from the withheld test set.

| Model Variant | Classification (CLS) |  | Regression (RG) |  |  | Survival (SRV) |  | Switching (SW) |  |
| --- | --- | --- | --- | --- | --- | --- | --- | --- | --- |
| | Accuracy↑ | AUC↑ | $R^2$ ↑ | RMSE ↓ | MAE ↓ | C-index<br>↑ | pAIC<br>↓ | Accuracy<br>↑ | AUC ↑ |
| M1<br>(Demographics + Clinical) | 0.77<br>(0.72,0.82) | 0.82<br>(0.77,0.86) | 0.65<br>(0.60,0.74) | 0.63<br>(0.55,0.69) | 0.43<br>(0.38,0.49) | 0.62 | 1905 | 0.97<br>(0.94,0.99) | 0.95<br>(0.88,0.99) |
| M2 (M1+Fluid Biomarkers) | 0.81<br>(0.77,0.85) | 0.83<br>(0.78,0.88) | 0.65<br>(0.57,0.72) | 0.64<br>(0.57,0.72) | 0.43<br>(0.38,0.48) | 0.65 | 1899 | 0.97<br>(0.95,0.99) | 0.93<br>(0.84,0.99) |
| M3 (M2 + Imaging Biomarkers) | 0.81<br>(0.77,0.85) | 0.85<br>(0.80,0.90) | 0.678<br>(0.62,0.76) | 0.61<br>(0.54,0.69) | 0.43<br>(0.38,0.48) | 0.68 | 1891 | 0.97<br>(0.95,0.99) | 0.94<br>(0.84,0.99) |

### 3.3 Longitudinal prediction of MCH incidence [RG & SRV]

We developed two types of longitudinal models for incidence of MCH over time in participants who were MCH-neg at baseline. The RG models were trained to predict the time (in years) to a participant’s first MCH-positive scan. The RG-M3 variant performed best, achieving a test-set R^2^ of 0.68 ± 0.079 and a RMSE of 0.61 ± 0.075 years (Table 2; Figure 2). For all three models, CDR-SB score was the primary predictor, with BMI and age as secondary and tertiary importance for RG-M1 and RG-M2, respectively (Figures 2d-2e). In RG-M3, the ICV-normalized gray matter and 1st Phenotypic Axis imaging score became second and fourth importance for predictive value, respectively, with BMI being third in importance (Figure 2c). Wilcoxon Signed-Rank tests confirmed a significant improvement for M3 over M1 (p=0.0005), and a marginal improvement for M3 over M2 (p=0.0064).

**Figure 2.** Longitudinal Prediction of Time to MCH Incidence (Regression Models). (A1–A3) Scatter plots illustrating the correlation between Predicted Duration (y-axis) and Actual Duration (x-axis) in years for the M3, M2, and M1 regression models. The dashed line represents perfect prediction (identity line). (B) Bar charts comparing R^2^, RMSE, and MAE across model variants with 95% confidence intervals. (C–E) Signed feature importance plots for each model variant.

The Cox-PH (SRV) models were developed to estimate the 48-month risk of MCH incidence. The SRV-M3 model demonstrated the best performance with a C-index of 0.68 and Partial AIC of 1789 (Table 2; Figure 3C). Across all SRV models, patient usage of oral diabetes medication was associated with the lowest log-hazard ratio for developing MCH, while patient clinical history of head injuries and usage of antidepressants was associated with the highest (Figure 3A). Kaplan-Meier survival analysis (Figure 3B) demonstrated robust risk stratification by the SRV-M3 model. The model successfully discriminated between risk tiers, with the High-Risk group exhibiting a markedly steeper decline in MCH-free survival compared to the Low-Risk group. While both medium and high-risk groups showed stepwise declines in survival probability at approximately 6 and 24 months, reflecting the protocol-driven intervals of MRI data collection in ADNI, the magnitude of these declines varied by predicted risk. By the 24-month mark, the high-risk group showed a significantly lower probability of remaining MCH-free compared to the low-risk group, confirming the model’s prognostic capability over the clinically relevant four-year follow-up.

**Figure 3.** Survival Analysis and Risk Stratification for MCH Incidence (Cox Proportional Hazards Models). **(A1–A3)** Forest plots displaying the Log Hazard Ratios (95% CI) for top predictors in the M3, M2, and M1 survival models. Positive coefficients (red) indicate increased risk of developing MCH, while negative coefficients (green) indicate reduced risk. **(B1–B3)** Kaplan-Meier survival curves stratified by model-predicted risk groups (Low, Medium, High). Shaded areas represent 95% confidence intervals. The models stratify MCH-free survival probability over a 4-year period. **(C)** Comparison of model fit using the Concordance Index (C-index, higher is better) and Partial AIC (lower is better).

### 3.4 Prediction of unstable MCH status [SW]

An exploratory analysis identified a sub-cohort of participants (∼15%) whose MCH status fluctuated between positive and negative across follow-up assessments. An XGBoost classifier was trained to predict which individuals would exhibit this “switching” behavior. No significant differences were found across M1, M2, M3, although SW-M1 achieved the highest AUC score of 0.96 ± 0.072 while retaining an accuracy of 97.6% ± 1.8%, which was shared with M2 and M3 (Table 2). Across all model variants, a clinical history of alcohol use, previous hepatic clinical history, and consumption of ophthalmologic medications were identified as important predictors of this unstable MCH phenotype.

## 4. DISCUSSION

This study introduces MCH-Guard, a multimodal machine learning framework designed to detect and predict spontaneous MCH in an observational cohort of cognitively unimpaired older adults and individuals with MCI or clinical diagnosis of dementia due to AD. Our analysis yielded three key findings. First, the framework detects the presence of MCH with high cross-sectional accuracy, even when utilizing only basic clinical data. Second, our longitudinal models predict future MCH onset, enabling patient-specific risk stratification over time. Third, we identified a novel transient vascular instability phenotype in approximately 15% of the cohort, challenging the conventional view of MCH as a strictly progressive pathology. Taken together, these findings demonstrate that a multimodal machine learning framework can provide a robust and clinically flexible decision-support tool for MCH risk management in the era of ATTs.

The high accuracy of our comprehensive cross-sectional classification model (CLS-M3, AUC 0.85) demonstrates the power of integrating diverse data modalities to identify patients with current MCH burden. The most predictive features, namely CSF p-tau181/Aβ42 ratio, plasma NfL, WMH, BMI, age, and cardiovascular history, align with the dual cascade hypothesis. This posits that both amyloid deposition in vessel walls (cerebral amyloid angiopathy) and tau-mediated neurodegeneration contribute synergistically to vascular fragility [10,21,28–30].

Notably, the strong predictive value of plasma NfL underscores the link between axonal injury and microvascular hemorrhage [40,41], a relationship further supported by the observed increase in spontaneous MCH incidence as the disease progresses over time (Supplementary Figure S2). While the inclusion of advanced imaging metrics—the unsupervised deep learning-derived T1-imaging phenotype (1st Phenotypic Axis score), normalized white matter hyperintensity volume, and normalized gray matter volume—yielded modest performance gains, these features likely capture complex, non-linear structural changes associated with AD and CAA progression that standard volumetrics miss.

Beyond single-time-point detection, our framework provides longitudinal prediction of future MCH development. The RG models explained a substantial portion of the variance in the time to MCH onset (R^2^ up to 0.68), with a MAE of approximately five months. Similarly, the SRV models effectively stratified patients into distinct risk tiers for developing MCH over a four-year period (C-index of 0.68). This predictive capability offers a method for creating patient-specific risk timelines, which could inform decisions for clinical trial enrollment and guide the frequency of safety monitoring for patients undergoing ATTs. For instance, individuals identified as longitudinally high risk could be scheduled for more frequent imaging to allow for early detection and treatment of spontaneous MCH & ARIA.

A particularly intriguing finding was the identification of a subpopulation (approximately 15%) exhibiting transient vascular instability—a phenotype characterized by MCH status fluctuating between positive and negative across visits. While often attributed to technical artifacts or hemosiderin resorption (‘vanishing microbleeds’) [42,43], our analysis suggests a systemic driver. Strong prediction of this instability by alcohol use and hepatic history (AUC >0.95) points toward a transient coagulopathy or reversible blood-brain barrier dysfunction associated with liver-brain axis interactions. This defines a distinct patient profile prone to vascular mimics that could confound safety endpoints in clinical trials, potentially leading to false-positive attributions of ARIA-H.

MCH-Guard integrates vascular, neurodegenerative, and systemic risk factors into a hierarchical framework adaptable to heterogeneous clinical settings. From a public health perspective, the minimal M1 model achieving an AUC of 0.82 using only demographics, clinical history, and basic cognitive scores indicates that effective risk stratification does not strictly require expensive fluid biomarkers or advanced neuroimaging. This has profound implications for global health equity and accessibility. The M1 variant of MCH-Guard could serve as a screening tool in primary care or resource-limited settings, identifying at-risk individuals who warrant referral for specialized imaging and fluid biomarker assessments, thereby democratizing access to safety assessments for disease-modifying therapies. Our MCH-Guard framework represents a significant advance over previous MCH risk-prediction efforts, which typically relied on single or categorical data modalities (e.g., only MRI, only APOE/demographics, or limited multi-imaging modalities), while requiring substantially fewer computational resources and less setup than MRI-based deep learning approaches [44,45]. By leveraging distinct data modalities, including CSF/plasma biomarkers, imaging, genetics, demographics, and comprehensive clinical patient information, we aim to capture the multifaceted nature of MCH pathophysiology, simultaneously incorporating vascular, neurodegenerative, and whole-body risk factors. We developed a hierarchical model variant framework to address the reality of heterogeneous data availability in clinical settings. Finally, we utilized T1 MRI latent embeddings derived from a self-supervised learning model to capture complex structural cerebral neurodegeneration, which may not be fully represented by traditional volumetric measures. Combined with other imaging metrics (ICV-normalized Gray Matter and ICV-normalized White Matter Hyperintensity), these latent embeddings contributed significantly to our comprehensive model.

Our findings have several important implications for the design and conduct of ATTs, particularly concerning patient selection, trial protocols, and personalized monitoring. The high-accuracy prediction of MCH risk allows for more nuanced inclusion/exclusion criteria. This precision could potentially expand access to life-changing therapies for patients currently excluded by blanket risk categories. For example, *APOE* ε4 carriers who have an otherwise low-risk profile based on our model might be considered for inclusion, provided appropriate monitoring protocols are in place. The identification of a small, but apparent MCH switching subgroup suggests that trial and MCH-related medication-screening protocols should account for potential spontaneous MCH variability when attributing imaging-derived symptoms to treatment effects. Further, this irregular switching behavior could also be problematic for the RG and SRV models, which rely on the typical developmental progression of MCH status. Incorporating serial baseline imaging before treatment, paired with patient demographics and medical history, may help identify individuals with inherent MCH instability. The strong cross-sectional and longitudinal predictive value of the CSF pTau181/Aβ42 ratio and plasma NfL suggests these biomarkers could serve as dynamic MCH risk indicators during treatment. Regular monitoring of these biomarkers, combined with our predictive models, could facilitate personalized dosing modulation strategies or enable early intervention for patients showing increasing risk profiles.

The minimal loss in predictive value between our minimal (M1) models and the more data-intensive (M2 and M3) models highlights the importance of baseline demographic and qualitative clinical data (e.g., clinical history, medications, neurological questionnaires) when initially evaluating a patient’s MCH risk.

Several limitations should be noted. First, our models were developed and validated using data on spontaneous MCH within the ADNI cohort. While the underlying pathophysiology is closely related to treatment-emergent ARIA-H, the models must undergo prospective validation in patient cohorts receiving ATTs before they can be implemented for risk stratification in the intended context. The baseline MCH burden is currently the strongest known predictor of ARIA-H in trials [46]. Therefore, predicting spontaneous MCH is the best available “proxy” for ARIA vulnerability in the absence of large-scale public trial data. Second, regarding the ‘switching’ MCH phenotype, we acknowledge that we cannot fully distinguish between biological resolution (hemosiderin resorption) and technical variability (lesions falling below detection thresholds due to head positioning or partial volume effects) without histological validation. However, the strong association of this phenotype with specific clinical predictors (alcohol, hepatic history) argues against this being purely random measurement noise. Further investigation is needed to clarify the basis of this observation. For simplicity and interpretability in the SRV modeling, we used only baseline biomarker and imaging values for training, though the use of dynamic values is acknowledged as a potential avenue for improving predictive value. Finally, we acknowledge that the ADNI cohort is predominantly Non-Hispanic White, which limits the immediate generalizability of these algorithms to minoritized populations. Given that vascular risk factors (e.g., hypertension, diabetes) have disproportionate prevalence in Black and Hispanic/Latino communities, validating MCH-Guard in diverse cohorts is a priority. Future work will focus on retraining these models within diverse datasets to ensure algorithmic fairness before clinical deployment.

In conclusion, MCH-Guard offers a data-driven approach to expanding patient access and improving safety in Alzheimer’s care. By integrating diverse data modalities within a flexible, tiered machine learning framework, our approach provides accurate cross-sectional detection and meaningful longitudinal prediction of MCH events. Its ability to deliver valuable risk stratification even with basic clinical data enhances its potential for broad and equitable clinical application. As ATTs become a cornerstone of AD care, tools like MCH-Guard will be essential for personalizing treatment strategies, maximizing therapeutic benefit, and minimizing safety risks, ultimately improving outcomes for patients with AD.

## Data Availability

All data produced are available online at adni.loni.usc.edu.

## ACKNOWLEDGMENTS

We would like to acknowledge ADNI for ADNI participant data analyzed in this study. Data collection and sharing for the Alzheimer’s Disease Neuroimaging Initiative (ADNI) is funded by the National Institute on Aging (National Institutes of Health Grant U19 AG024904). The grantee organization is the Northern California Institute for Research and Education. In the past, ADNI has also received funding from the National Institute of Biomedical Imaging and Bioengineering, the Canadian Institutes of Health Research, and private sector contributions through the Foundation for the National Institutes of Health (FNIH) including generous contributions from the following: AbbVie, Alzheimer’s Association; Alzheimer’s Drug Discovery Foundation; Araclon Biotech; BioClinica, Inc.; Biogen; Bristol-Myers Squibb Company; CereSpir, Inc.; Cogstate; Eisai Inc.; Elan Pharmaceuticals, Inc.; Eli Lilly and Company; EuroImmun; F. Hoffmann-La Roche Ltd and its affiliated company Genentech, Inc.; Fujirebio; GE Healthcare; IXICO Ltd.; Janssen Alzheimer Immunotherapy Research & Development, LLC.; Johnson & Johnson Pharmaceutical Research & Development LLC.; Lumosity; Lundbeck; Merck & Co., Inc.; Meso Scale Diagnostics, LLC.; NeuroRx Research; Neurotrack Technologies; Novartis Pharmaceuticals Corporation; Pfizer Inc.; Piramal Imaging; Servier; Takeda Pharmaceutical Company Limited; and Transition Therapeutics.

## ADNI Collaborators

Data used in the preparation of this article were obtained from the ADNI database ([7]). As such, the investigators within the ADNI contributed to the design and implementation of ADNI and/or provided data but did not participate in the analysis or writing of this report. A complete listing of ADNI investigators can be found at: http://adni.loni.usc.edu/wp-content/uploads/how_to_apply/ADNI_Acknowledgement_List.pdf

## CONFLICTS

A.P., E.P., I.H., P.M. have nothing to disclose. D.T. reported grants from National Institutes of Health during the conduct of the study.

## FUNDING SOURCE

Supported in part by the National Institutes of Health (grant U01AG024904)

## CONSENT STATEMENT

Data collection and sharing for this project was authorized by the Alzheimer’s Disease Neuroimaging Initiative (ADNI). ADNI participants provided written informed consent at the time of enrollment for data collection and completed questionnaires approved by each participating site’s Institutional Review Board (IRB).

## Notes

### Competing Interest Statement

The authors have declared no competing interest.

